# Co-production of an oral health intervention: Enabling Service Users with Severe Mental Illness to Learn about and Engage (SMILE) with Good Oral Health

**DOI:** 10.64898/2026.11.06.26367413

**Authors:** MP Mishu, Natalia Kika, Emma Tkach, James Bird, Jade Cupac, Sarah Daniel, Michelle Horspool, Gordon Johnston, Helen Killaspy, Zoe Marshman, Louise Marston, Sadia Nafees, Siyabonga Ndwandwe, Shengning Pan, Eshan Vadgama, Florence Watson, Emily Peckham

## Abstract

**Background:** People with severe mental illness (SMI) experience poorer oral health than the general population. They are less likely to visit the dentist or brush their teeth and face unique barriers to accessing oral health support. There is a limited number of oral health interventions for people with SMI, and there is no conclusive evidence on their effectiveness. An integrated oral health intervention delivered by mental health professionals in a community mental healthcare setting could offer a scalable option to oral health support for people with SMI. In this paper we report on the process of co-producing such an intervention in England.

**Methods:** The intervention development process was informed by Hawkins’ co-production framework. In this paper we report on stages 2 (co-production) and 3 (prototyping). Two online and two face-to-face co-production workshops took place (N=22) involving service users with SMI (n=6), mental health professionals (n=8) and dental health professionals (n=8). Two online patient and public involvement consultations, and a professional stakeholder meeting were held during the co-production process to provide feedback on the co-production process. The workshops informed the core components, format and content of the intervention.

**Results:** The co-production process resulted in five intervention components: 1) Initiating a conversation about oral health, 2) providing advice to people with SMI on keeping their teeth and mouth healthy, 3) weekly tooth brushing text reminders, and 4) mental health professionals updating the care record to document the administration of this intervention. To support intervention delivery, we co-produced an intervention flowchart, an intervention leaflet for people with SMI, and a training manual for mental health professionals delivering the intervention.

**Conclusion:** We co-produced an integrated oral health intervention to be delivered to service users with SMI in community mental health settings. The next stage of the research is a single-arm feasibility study to assess whether the intervention is acceptable to people with SMI and feasible to deliver within the NHS.

## Background

Oral health is an integral part of health and wellbeing. Oral diseases are a common cause of pain, infection and tooth loss, leading to functional limitations such as eating difficulties which can affect nutrition (1). Poor oral health also has negative psychosocial impacts on self-esteem, social interaction, and overall quality of life and is related to other systematic diseases such as diabetes and cardiovascular disease (1). Oral diseases are a global public health concern with a significant economic burden. In 2019, UK households’ expenditure on dental services was more than £5.96 billion (2).

Oral health among people with severe mental illness (SMI) is poorer than that of the general population. They are nearly three times more likely to lose all of their teeth (OR = 2.81, 95% CI [1.7 -4.6]), have higher rates of dental caries (mean difference 5.0, 95% CI 2.5 -7.4) and are less likely to visit a dentist for routine dental check-ups or treatment (3). These poor outcomes are due to several factors (4) including symptoms of mental illness being associated with decreased motivation for regular tooth brushing (5), high consumption of sugary food and drinks (6), and side effects of medications (e.g. dry mouth) (5).

Support with the maintenance of good oral health and attending regular dental check-ups is scarce for people with SMI, which only exacerbates the poorer oral health outcomes among this population. This issue also contributes to serious complications such as hospital admissions due to physical health complications from untreated dental decay (7), accompanied by associated resource implications for the health service. People with SMI also often require specialist dental care and general anaesthesia, partly due to seeking treatment late.

However, to improve oral health, people with SMI require interventions that are tailored to their needs. This is crucial given that this population faces both individual barriers, such as a lack of trust in dental health professionals, as well as mental health symptoms; and organisational level barriers, including limited staff capacity and resources to address their dental health concerns. Furthermore, these complex needs and barriers to oral health support among people with SMI require an approach that integrates oral health care into mental health services where possible. This need is also underlined by the calls for approaches which integrate physical and mental healthcare to reduce health inequalities (8). To synthesise and evaluate currently available oral health interventions for people with SMI, we previously conducted a systematic review and meta-analysis which identified 12 studies (9). We found that the interventions covered five broad categories: dental education, motivational interviewing, dental checklists, dietary change and incentives. Although there were some statistically significant improvements in dental plaque index and oral health-related behaviours, we found that the studies included in the review were of low to moderate quality; thus, the clinical effectiveness of oral health interventions for people with SMI remains unclear. Only one intervention was delivered by mental health professionals in a UK community mental health setting; no effects on service users’ oral health outcomes were found after a one-year follow-up (10). Additionally, none of the interventions were co-produced or reported public and patient (PPI) involvement, meaning that the voices of service users, their carers, and healthcare professionals were not necessarily reflected in the respective intervention development processes. Given that co-production is a key aspect of evidence-based intervention development guidance (11), and the unique challenges of service users with SMI, it is crucial to involve them in the development of any future oral health interventions.

To explore potential avenues of addressing the limitations of existing interventions, we began the process of co-producing an intervention to improve oral health in people with SMI. This was informed by Hawkins’ co-production framework which aims to provide guidance on co-producing and prototyping intervention materials with relevant stakeholders before proceeding with their evaluation (12). The framework therefore involves three stages: 1. Evidence review and stakeholder consultation, 2. Co-production and 3. Prototyping. Stage 1 included our systematic review on oral health interventions for people with SMI, and initial stakeholder consultations to identify preliminary key ingredients for a new oral health intervention (13). Here we conducted consultations with 23 stakeholders including service users, carers, dental and mental health professionals, and dental public health professionals. The potential intervention steps we identified were: 1) initiating the conversation around oral health; 2) delivering tailored oral health advice; 3) providing encouragement to reinforce the delivered advice; 4) including oral health in the service user’s care plan; 5) facilitating visits to the dentist. More detail about the involved stakeholders and the process of identifying these intervention steps is reported elsewhere (13).

Having completed Stage 1 and identified the preliminary core components of an oral health intervention, the next step was to co-produce an intervention (Stage 2) with accompanying training resources and prototyping the intervention (Stage 3), Stages 2 and 3 are reported in this article (Figure 1). The ultimate aim of this co-produced intervention is to improve oral health in people with SMI.

**Figure 1:**
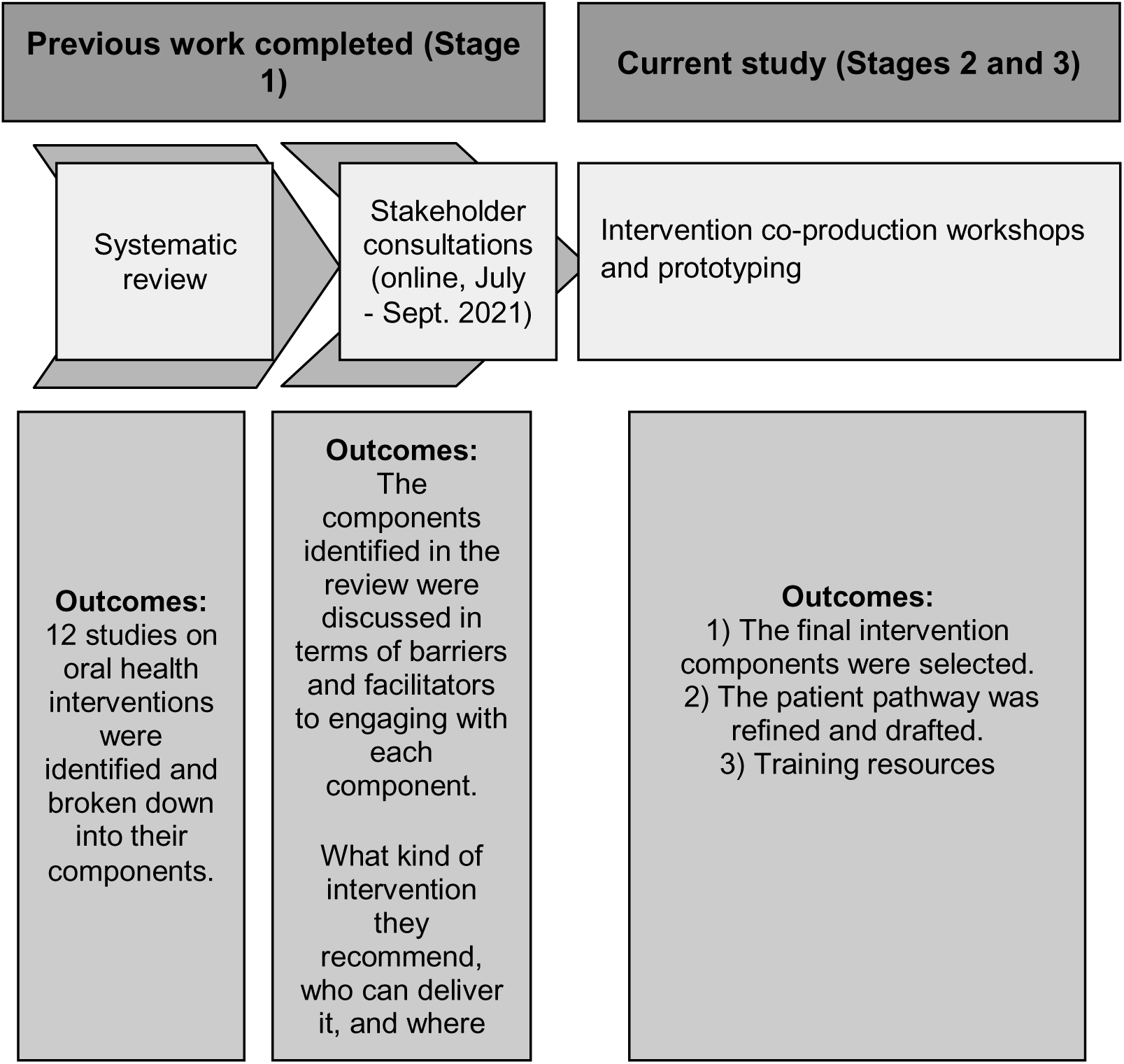
Previous and current work to co-produce this intervention

### Aims and Objectives

In this paper we describe the process of co-producing an intervention to improve oral health in people with SMI that is acceptable to service users and feasible to deliver within routine mental health care. This was one component of the SMILE project which also aims to test the feasibility of the co-produced intervention (the feasibility study findings will be reported elsewhere). The co-production component of the project had two objectives:

1. To develop, through co-production with people with SMI, caregivers, service providers and related stakeholders, an intervention to improve oral health and oral health quality of life and provide support to access dental services.
2. To co-produce intervention materials, including i). an intervention leaflet for service users; ii.) a training manual and training materials for staff, including an intervention flowchart.

## Methods

### Study design

We drew on Hawkins’ (2017) three-stage intervention co-production model, based on recommendations for co-production approaches to intervention development whereby stakeholders are actively involved across planning, developing, and testing of interventions (12). As described in the background section, we previously completed Stage 1 of Hawkins’ model, comprising an evidence review and stakeholder consultations to establish the initial intervention components. In this paper we report on stages 2 (co-production) and 3 (prototyping) to refine the intervention and produce its related outputs. The intervention development process is reported in line with the GUIDED checklist (14)(Additional File 1), and the intervention content followed TidiER guidance to describing interventions (15).

#### Ethics

We obtained a favourable NHS ethical approval from the West Midlands – South Birmingham Research Ethics Committee for the co-production workshop component of the study (ref. no, 24/WM/0267).

#### Recruitment

We recruited participants with lived experience of severe mental health, dental, and mental healthcare professionals to take part in intervention co-production workshops. Recruitment channels included professional networks, including mental and dental healthcare colleagues and the two NHS trust sites involved with the SMILE project. All participants had to be aged 18 or over, have capacity to consent to participation and be willing to be audio recorded during the workshop. Additional eligibility criteria that applied to service users were having a verified diagnosis of an SMI such as schizophrenia, schizoaffective disorder, bipolar disorder or other non-organic psychosis, carers had to identify as supporting someone with an SMI, and healthcare professionals included mental health professionals providing support to people with SMI or professionals involved in the delivery of oral health care. Some PPIE group members also took part in the co-production workshops as participants, providing a link between the PPIE group and the workshops.

#### Procedure

To co-produce the intervention, we carried out four co-production workshops described below which were based on individual topic guides for each workshop (Additional File 2). The formal co-production process was accompanied by two PPIE meetings with people with SMI and their carers, and one meeting with mental health professionals. The two stakeholder groups were consulted separately to provide ongoing feedback on the iterative intervention development process and to inform workshop planning.

#### PPI group meetings

Two online PPIE meetings with service users were held alongside the workshops to guide the iterative process of refining the intervention (Figure 2). The meetings were attended by a total of nine service users and were conducted after the first and second workshop, respectively. Both meetings focused on refining the chosen outcome measures for the feasibility trial, the steps involved in delivering the intervention and aspects of oral health-related advice agreed on in the workshops. Input from the PPIE meetings directly informed the structure and topics of subsequent co-production workshops. Before the first workshop we held two PPIE meetings with n=3 service users, and n=2 mental health professionals in total. We held an additional mental health professional PPIE meeting after the final workshop (n=4). This was to seek guidance from this group on which suggested changes from the final workshop would be feasible to implement.

**Figure 2.**
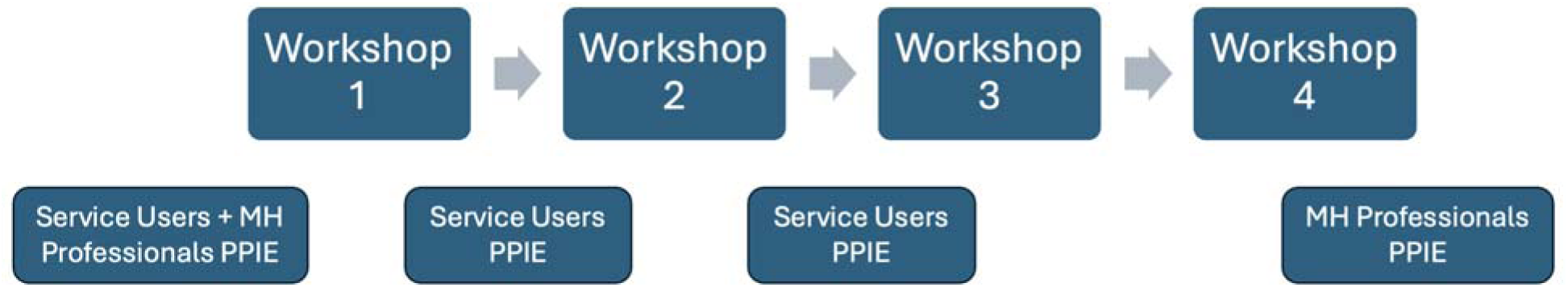
The intervention co-production process with concurrent PPIE meetings.

### Co-production (Stage 2)

#### Workshop 1

The first workshop was conducted online on MS Teams to maximise accessibility and reduce geographical barriers to participation. Participants were service users with SMI (n=3), a carer (n=1), mental health professionals (n=4), dental health professionals (n=6). The workshop was facilitated by three research team members, one of whom is also a dental health professional. Here we presented the five-step intervention structure which was developed based on the initial stakeholder consultations from Stage 1. The initial five steps were: 1) mental health professionals initiating the conversation around oral health with the service user; 2) delivering tailored oral health advice; 3) providing encouragement to maintain good oral health habits; 4) including oral healthcare in the service user’s care plan; 5) providing help to support the service user to visit a dentist if needed. Research team members facilitated in-depth discussions for each intervention step in two breakout rooms, ensuring a balance of various stakeholders’ perspectives in both rooms.

We received detailed input across the initial intervention steps. Participants discussed the various ways in which a conversation about oral health could be started, and all agreed on using non-stigmatising language to avoid causing feelings of shame and embarrassment among service users. The discussion included details on where the oral health conversation would take place; it was agreed that it was most fitting to incorporate it into routine conversations around physical health, rather than a standalone conversation which could make service users self-conscious about their oral health. When giving practical advice on maintaining good oral health (e.g. toothbrushing demonstration, dietary sugar intake, tobacco and alcohol), it was important that people delivering the intervention used an encouraging tone to avoid sounding patronising. Suggested approaches included asking service users for permission to receive the advice, e.g. ‘Would you like to see a toothbrushing demonstration?’, rather than imposing the advice. Participants suggested that people delivering the intervention should clearly explain the links between the importance of lifestyle habits (diet, tobacco and alcohol) and their impact on oral health specifically, rather than just the better-known links with general health.

#### Workshop 2

The main focus of the second workshop (online) was to discuss which outcome measures would be suitable for the upcoming feasibility study (Mishu et al., under review) to test the co-produced intervention. Participants were service users (N=2), a carer (N=1), mental health professionals (N=3), and dental health professionals (N=2). A summary of the discussion on outcome measures is provided here.

Emphasis was placed on simplifying the initially proposed outcome measures such as dental plaque index, and detailed measures of toothbrushing habits, dietary sugar intake, alcohol and tobacco consumption. Dental and mental health professionals agreed that collecting clinical outcome measures of oral health indicators were beyond the scope of mental health professionals. Service users felt that taking such measures using clinical instruments could be intimidating. Mental health professionals and service users suggested that asking for precise habit data may lead to recall bias or inaccurate information recall. The agreed measures which were deemed acceptable for a further feasibility evaluation covered information on oral health status, including broad toothbrushing habits, dental visits, dietary sugar intake, alcohol and tobacco use, a custom resource use questionnaire refined with stakeholders, and standardised outcome measures – oral health and health-related quality of life.

#### Workshop 3

The third workshop was conducted face-to-face. Participants were people with lived experience of SMI (n=2), one carer, mental health professionals (n=3), dental health professionals (n=4), and a health economist (n=1). Three researchers facilitated the workshop. Based on the findings from the first workshop, we developed a topic guide in which we focused the discussion on the second intervention component -delivering oral health advice, and the four advice categories within it: 1) oral hygiene; 2) dietary sugar intake; 3) tobacco and alcohol consumption; 4) side effects of psychotropic medication. We also discussed the practicalities of delivering the intervention, including who was best placed to initiate the conversation, and how the initial question should be phrased. We then sought participants’ input on the images used in the intervention materials to ensure they were acceptable and relevant to the target population.

Following the first workshop we refined the initial intervention steps and drafted a document outlining a skeleton of the SMILE intervention leaflet. This draft version contained an overview of the intervention steps, as well as preliminary images and graphics which were to be developed further. Participants suggested using an image to facilitate initiating the conversation, which would be through the link between general and oral health.

While service users expressed the need for support with visiting a dentist, healthcare professionals agreed that it would not be feasible for people delivering the intervention or care co-ordinators to actively look for dentists for service users due to time and capacity constraints. Therefore, we decided to include a step on encouraging and optionally supporting the service user to visit a dentist, should the person delivering the intervention have the capacity to do so.

#### Workshop 4

The fourth and final workshop was conducted face-to face. Here we presented the revised intervention components based on participants’ previous input from workshops 1 and 3 and continued to discuss and refine any outstanding issues or areas of uncertainty. We also focused on the intervention flowchart to discuss the order in which the intervention components would be delivered. Participants were people with lived experience of SMI (n=3), one carer, mental health professionals (n=3), and dental health professionals (n=4). Four researchers facilitated the workshop in two breakout groups.

In this workshop we finalised the content of the intervention and leaflet, and the order in which the steps would be delivered. Participants suggested decreasing the amount of text to make it easier to follow. Participants agreed on which images were to be adapted by a graphic designer for the leaflet, and suggested modifications such as reductions in the amount of text in the images. We also made some changes to the wording based on service users’ suggestions, e.g. using ‘top advice’ instead of ‘top tips’. An issue regarding the intervention flowchart was that conversations with service users would not always follow a linear structure in real-life intervention delivery settings. Therefore, we decided to emphasise this fluidity of intervention step delivery to staff delivering the intervention in any subsequent intervention materials. We also agreed to remove the provision of a toothbrushing demonstration, as mental health professionals advocated that they would not feel comfortable providing this, given that it is outside their scope of practice. Therefore, we opted for a graphic to illustrate this in the leaflet with accompanying text instructions and a link/QR code to a video demonstration. To ensure the tailored element to the intervention, we added free-text space under each intervention step where service users could note down their own barriers and goals relating to each step (e.g. toothbrushing -what is currently getting in the way of regular toothbrushing?).

A key change introduced following the final workshop was that mental health professionals felt 1) the inclusion of the intervention in the service users’ care plan, and 2) support with visits to a dentist or oral health professional, would not be implementable, as part of a feasibility study, due to staffing resources. Therefore, a compromise was agreed whereby those delivering the intervention were to record in the service users’ care records the details of their personal oral health intervention plan and any concerns or advice given by oral health professionals or dentists where such visits had occurred as part of the intervention. The proposed step of mental health professionals supporting service users with visits to oral and dental health professionals was removed from the intervention.

The final intervention components as agreed after the final workshops were as follows: 1) Initiating a conversation about oral health; 2) providing advice to service users on keeping their teeth and mouth healthy; 3) weekly text reminders for tooth brushing; and 4) updating the service user’s care record to document the details of the intervention plan. Table 1 offers a summary of the key changes made to the initial intervention components based on stakeholders’ input.

**Table 1.** Initial intervention components, key stakeholder input and its resulting intervention content changes.

| Initial intervention component | Stakeholder input | Implemented changes |
|---|---|---|
| Mental health professional initiates the conversation around mental health | Arranging separate conversations about oral health may be uncomfortable and make service users feel self-conscious – a way to overcome this is to include it into regular appointments | Conversation embedded into existing appointments, e.g. physical health checks |
| Providing tailored oral health advice on:<br>1) Toothbrushing<br>2) Dietary sugar intake<br>3) Dry mouth<br>4) Tobacco<br>5) Alcohol | 1) - Toothbrushing – exclude provision of toothbrushing model (out of scope for mental health professionals), Provide QR code/link to demonstration video + illustrations in the leaflet<br>- Tiered approach to tailor advice to current toothbrushing frequency | Removed the provision of a toothbrushing demonstration using a model; illustration of toothbrushing technique and a video QR code included in the intervention leaflet |
|  | 2) Dietary sugar intake:<br>- Focus on what should be added, rather than removed from one's diet<br>- Realistic suggestions – sugar-free swaps instead of restrictions | Refined advice to a more realistic approach – changed phrasing to reflect healthier swaps rather than dietary restrictions, considering service users' struggles with craving due to medication side effects |
|  | 3) -Dry mouth – advice should be brief and general, focus on dry mouth management without emphasising it is a side effect, to avoid decreasing adherence to medication<br>-Avoid recommending products which require purchases | The dry mouth advice component is brief and provides general advice on using water or chewing sugar-free gum to alleviate symptoms.<br>Products such as artificial saliva sprays are not included in the advice. |
|  | 4) Tobacco and alcohol:<br>Advice is already routinely provided in community mental health service; only cover briefly in the SMILE intervention | Advice on tobacco and alcohol is brief, general, and focuses on how their consumption affects oral health<br>Signposting to support with tobacco and alcohol included in the leaflet |
| Provide encouragement to maintain good oral health habits | Text message and phone call check-ins were discussed as an approach to positive reinforcement of toothbrushing habits | One text message toothbrushing reminder sent by mental health professional; same message content to be shared with all service users |
| Include intervention in the care plan | Service users supported this step so that oral health advice and action points are followed up on<br>Mental health professionals expressed concerns due to care coordinator capacity | The intervention plan is recorded in the service user's care record, rather than care plan, so that care co-ordinator follow-up is encouraged but not mandatory |
| Provide support with visiting a dentist | Stakeholders discussed the possibility of care coordinators identifying available dentist practices and supporting service users' visits<br>Mental health professionals expressed concerns due to care coordinator capacity<br>Dental professionals expressed concerns about dental practice availability and variation across regions | This component was removed as a standalone step – if the service user does not have a dentist, this will be recorded in their care record and included in a letter to their care coordinator; assistance with dental visits will be encouraged but optional based on staff capacity. |

## Results

### Prototyping (Stage 3)

Following the final workshop, we drafted the intervention pathway, intervention leaflet (Additional File 3) and training manual for the mental health care professionals and a SMILE intervention flipchart to be used by the mental health professionals when delivering the intervention. We followed a thorough iterative process for co-developing these, whereby drafts were refined and re-visited on an ongoing basis with feedback from co-investigators, service users and carers, mental and dental health professionals and an external graphic designer until the final versions were agreed.

To ensure thoroughness during the iterative process, this step took three months longer than we expected. However, we did not want to compromise the co-design element of the intervention and training resources, and therefore we decided to extend the timeline for this purpose.

Three main intervention outputs were co-produced: the participant-facing intervention leaflet’ the intervention flowchart; and the training manual for interventionists – mental health professionals who will be delivering the intervention.

### SMILE Intervention

The final intervention prototype consisted of the following components:

**1. Initiating the conversation:** the health professional delivering the intervention opens the conversation by highlighting the link between oral and general health, referring to a corresponding illustration in the leaflet. Additional prompts such as ‘Do you feel you are satisfied with your oral health?’ are offered to encourage a discussion. Service users with urgent dental health problems are advised to call 111.
**2. Top advice to keep your teeth and mouth healthy:** this is the main section of the intervention, covering advice on four areas of habits related to good oral health 1) tooth brushing, including advice on good technique through an illustration, a video link, and written instructions. Service users can note down their current toothbrushing habits and tooth brushing frequency goals based on a tiered system to tailor the recommendations offered to them, depending on their level of readiness to engage with the intervention; 2) managing dry mouth due to the side effects of psychotropic medication; 3) reducing dietary sugar intake through food and drinks -service users can note down their current barriers to reducing their intake; 4) stopping or reducing tobacco use, if ready and willing to, with advice on local cessation support services; 5) responsible alcohol drinking with NHS advice on weekly alcohol unit consumption and information on local alcohol support services; 6) visiting a dentist, explaining the importance of regular dental visits and discussing individual barriers to attending dental appointments.
**3. Sending reminder text messages:** Service users can opt into receiving a weekly standardised text message reminder to encourage regular tooth brushing for the first four weeks from when the intervention delivery has commenced.
**4. Recording the intervention in the care record:** A note is added to the service user’s care record to document that they have received the SMILE intervention. If the health professional delivering the intervention, carer, or service user raises any concerns about their oral health, or barriers to receiving dental health support, these are also recorded here so that their mental health team are aware of them. A letter is sent to the service user’s care co-ordinator to confirm the same information and bring their attention to any urgent dental health issues to ensure follow-up.

### Intervention materials

#### Flowchart

The intervention flowchart (Figure 3) presents the structure and intervention steps: 1) initiating the conversations, 2) providing key advice on maintaining good oral health, 3) sending a weekly toothbrushing reminder for four weeks, 4) documenting the intervention in the service user’s electronic healthcare record and informing their care co-ordinator. The flowchart further illustrates the options to tailor intervention step 1 and 2 based on the service user’s preferences and current oral health support needs.

**Figure 3.**
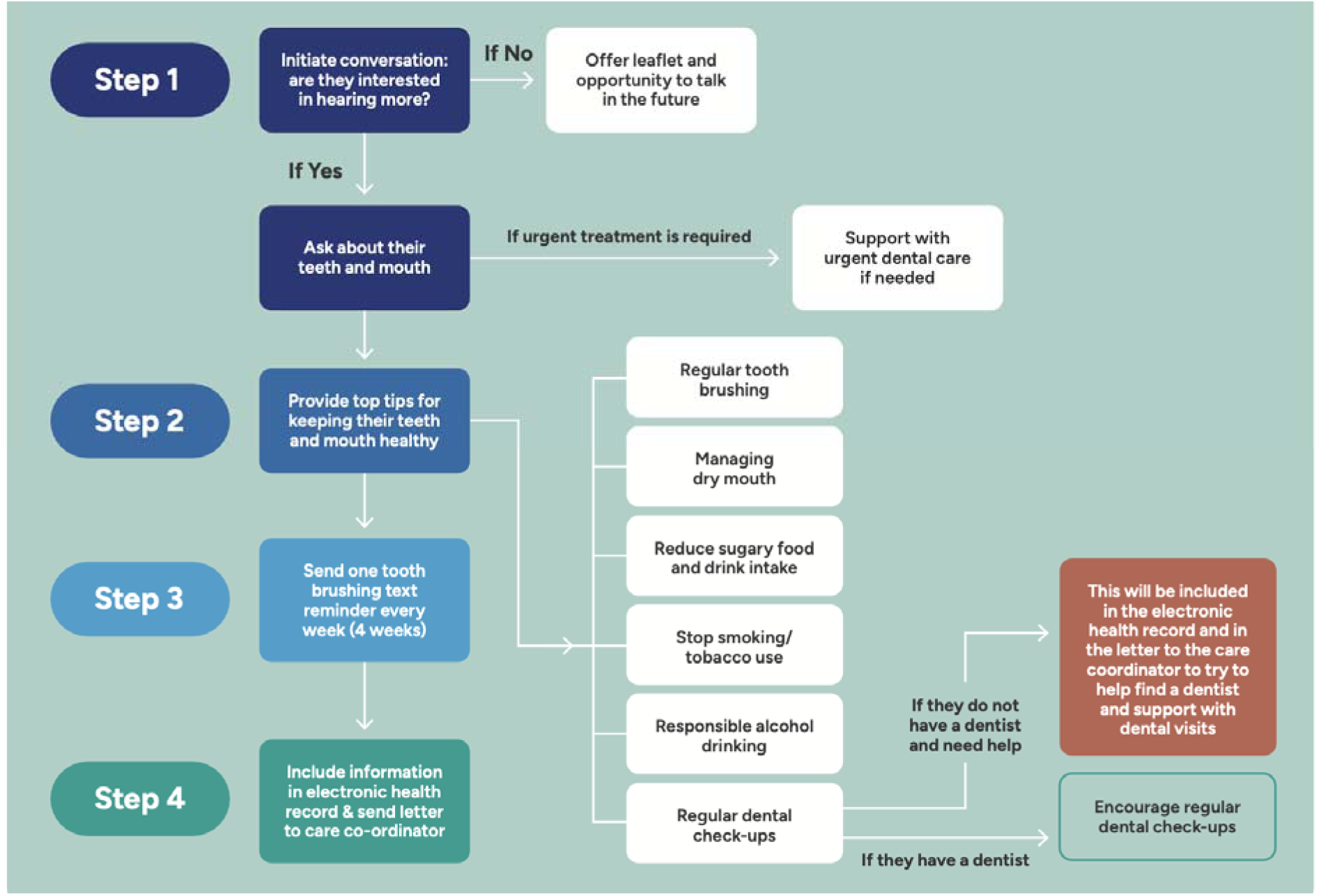
SMILE intervention flowchart.

#### Intervention Leaflet

Following the participants’ input on the overall structure and outline of the leaflet content, the research team developed the detailed leaflet content (Figure 4). This step was conducted iteratively, with written and verbal input until a final version was agreed upon with service users and carers and research team. This was then developed and refined with a graphic designer, with the researchers sharing participants’ feedback when reviewing the designer’s drafts. The research team and PPIE group were generally satisfied with the leaflet; minor changes were suggested and made, relating to the choice of wording, reducing the amount of text to make the content easier to read, and including signposting information for tobacco and alcohol services.

**Figure 4.**
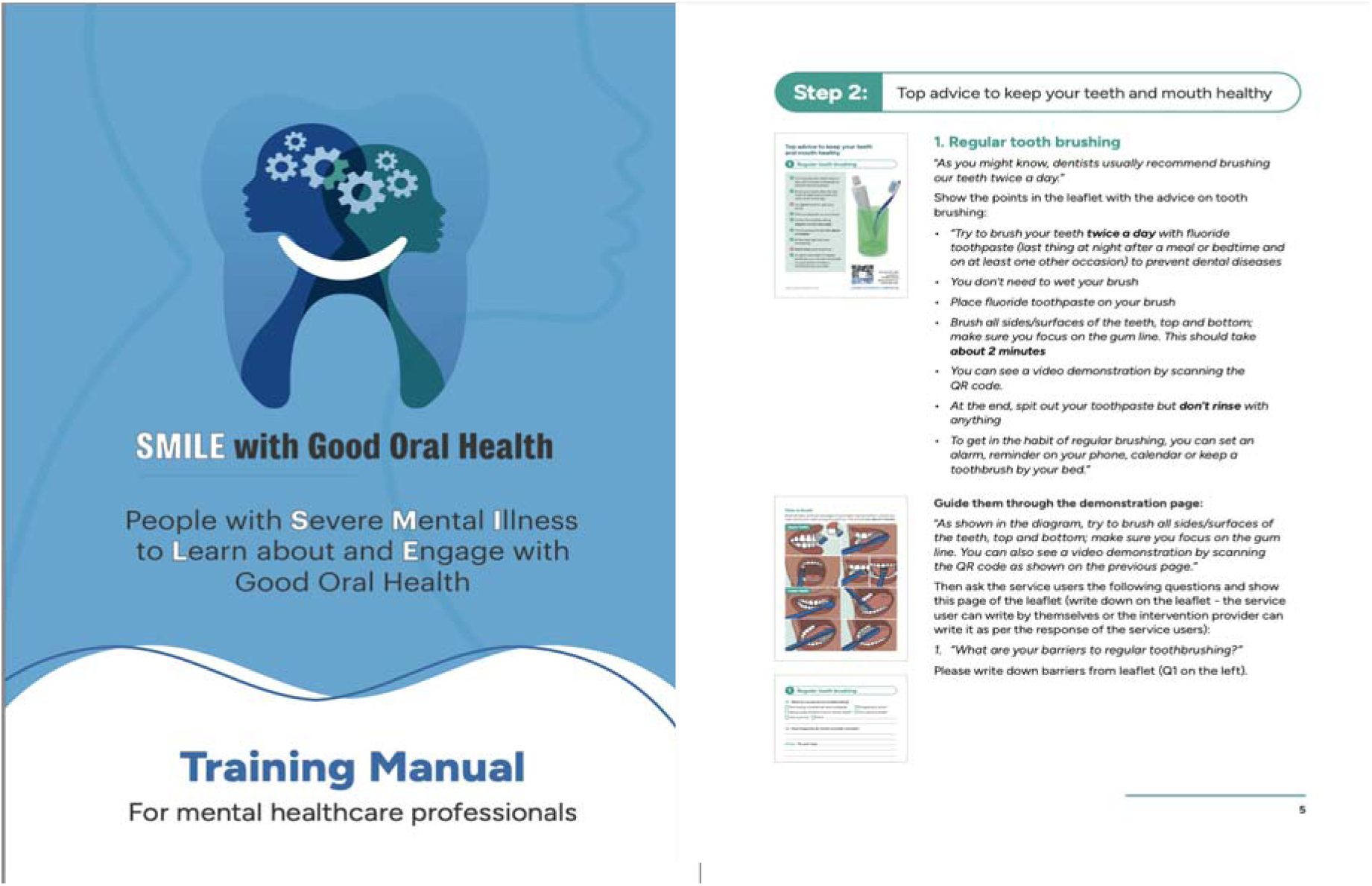
SMILE intervention leaflet examples.

#### Training Manual

The training manual (Fig.4) provides the background and aims of the SMILE interventions to the mental healthcare professional delivering it. It follows the SMILE intervention stages by providing steps to delivering each stage of the intervention. Guidance focuses on optional questions interventionists can ask to start the conversation around oral health, as well as steps to ask about the service user’s individual challenges with implementing oral health related lifestyle habits.

## Discussion

This paper describes the process of co-production used to develop the SMILE intervention, a novel intervention to offer oral health advice to community mental health service users. Through an iterative co-production process consisting of four co-production workshops, concurrent PPIE meetings, and prototyping iterations with stakeholders, we developed an intervention consisting of four steps: 1) mental health professionals initiating a conversation with service users around oral health, 2) mental health professionals providing top advice to service users on keeping their teeth and mouth healthy, 3) mental health professionals sending weekly text reminders for toothbrushing and 4) mental health professionals updating the care record to document the administration of this intervention. We co-produced three intervention components to support intervention delivery: 1) intervention flowchart, 2) intervention leaflet, and 3) training manual for mental health professionals. The overall structure of the intervention remained similar to the initial proposed intervention steps; however, several amendments and considerations were suggested during the iterative process of producing the intervention components.

Suggestions were made by participants to simplify the intervention content to avoid overwhelming service users with advice. The benefit of keeping oral health advice simple was considered to be greater than providing more comprehensive and lengthy oral advice; as such, habitual changes were often difficult to implement and the success of service users implementing one change was deemed more motivational and sustainable than attempting to implement many, with perhaps varying levels of success. This was further understood and substantiated through the tiered approach which we used to understand the ‘readiness’ and capacity of service users to engage with the intervention.

Mental health professional participants advised that we reduce the tailored aspect of the intervention to ensure the advice they were being asked to give service users on their oral health was within the scope of their expertise. The overarching intention of the SMILE intervention was to increase accessibility of oral health information and dental hygiene advice amongst service users with SMI. To do so, the intervention must be deemed usable and understandable by the mental health professionals tasked with delivering it. Therefore, we removed the originally proposed toothbrushing model demonstration, and scaled back the recording of the intervention from the service users’ care plan to their care record, as a recommendation for care co-ordinators to support dentist visits where needed.

Our previous work on understanding the barriers to oral health access for people with SMI identified the impact of mental ill-health itself, lack of patient involvement, and a lack of accessibility and availability of dental services for this group. Barriers contributing to oral health inequities amongst people with SMI compared to the general population also included a low level of integration of dental and mental health services, and limited inter-disciplinary working. The main facilitators of better access to oral health were more effective communication skills of service providers to service users, and further support through the involvement of carers (4).

The co-production of the SMILE intervention components (i.e. the intervention flowchart, intervention leaflet, and the training manual for interventionists) specifically aim to reduce the barriers and utilise the facilitators identified in this previous work.

As identified by a recent systematic review and meta-analysis on mental providers’ attitudes, knowledge and practice relating to providing oral health support (16), there is unexplored potential for mental health professionals supporting service users with their oral health. Our intervention addresses this gap; importantly, by co-producing SMILE with key professional and service user stakeholder groups, we sought to ensure the intervention is simple to administer with minimal time resources, given the commonly identified workload challenges among mental health professionals (17).

Furthermore, this study aligns with Palmier-Claus’ et al. ’s (18) seminal randomized-control trial (RCT) assessing the impact of link-workers in supporting service users with SMI in accessing dental care and services, with the aim of improving their oral health. Link workers were support workers within NHS mental health services, and they were specifically tasked with providing support for people with SMI in attending a routine dental appointment during the intervention. Findings from this study were promising in that the intervention was deemed acceptable, feasible and safe, and initial results showed promise in terms of clinical effectiveness. Our study adds to such findings in that it provides a framework for understanding the process behind co-production of such an intervention, which engages stakeholders ranging from those receiving the care, and those implementing/delivering the care. Such insights are beneficial in modifying and developing the intervention which, if found to be effective, may eventually become integrated into standard care provision.

### Strengths and limitations

The approach taken in this study presents several strengths in this field of research. Firstly, the tiered approach adopted during co-production was an important strength for developing an effective and practical intervention for service users with SMI. Given the range of complex needs that service users with SMI experience, such an approach is important as it bases evaluations of intervention components through hierarchical levels of ‘readiness’ and capacity of individuals to engage with the intervention. Secondly, this intervention signifies important progress in the field of oral health of people with SMI: the SMILE intervention is a novel system-level approach to both empowering service users to engage with preventive action to maintain and improve their oral health, while also working with MHPs and existing healthcare settings to provide systemic support for people with SMI. This builds upon our previous work which found that barriers to receiving dental care for people with SMI were generally cross-cutting across personal, interpersonal and system-levels (4) and specifically addresses both bottom-up and top-down approaches to improving the oral health of service users with SMI.

However, this study also presents with several limitations. Firstly, we recognise the lack of diversity in the stakeholder representation from different ethnic backgrounds and non-English-speaking groups. While we aimed to transparently report on a pragmatic, iterative intervention development approach with ongoing real-time feedback and refinements from multiple stakeholder groups, this also meant that we did not conduct formal qualitative analysis of workshop data. Therefore, it is possible that a more systemic, in-depth understanding of stakeholders’ needs and preferences had been overlooked at times.

### Future directions

The next step is to conduct a planned single-arm feasibility trial of the co-produced intervention via two NHS sites. If the intervention proves to be feasible and acceptable to service users and mental health professionals, a larger-scale pilot study with a control group would be the next step. If the current study indicates the SMILE intervention could be feasible and acceptable, additional professional stakeholder consultations could include a broader range of stakeholder backgrounds, such as health commissioners and health economists, to explore how the intervention could be implemented at the wider system level. We realise the need to engage with broader groups of stakeholders to understand the acceptability and feasibility of integrating this intervention within a public healthcare system like the NHS.

### Conclusions

In this paper we described the co-production process of SMILE, a novel systems-level oral health intervention for community mental health service users with SMI. We shed light on the needs and preferences of service users, mental and dental health professionals, and showcased an example of how an intervention can be developed pragmatically with real-time engagement and refinements from the target user group and professional groups involved in intervention delivery and content. The next step for the SMILE intervention will be to conduct feasibility and acceptability testing to evaluate whether the intervention can progress onto further effectiveness testing.

## Data Availability

The intervention leaflet is attached to this manuscript (Additional File 3). Summaries of co-production workshop discussions are available upon reasonable request.

## List of abbreviations

NHS: National Health Service
PPIE: public and patient engagement
SMI: severe mental illness
SMILE: Severe Mental Illness to Learn about and Engage with Good Oral Health

## Declarations

### Ethics approval and consent to participate

We obtained a favourable NHS ethical approval from the West Midlands – South Birmingham Research Ethics Committee for the co-production workshop component of the study (ref. no, 24/WM/0267), in accordance with the Declaration of Helsinki.

### Consent to Participate

All participants provided written informed consent to take part.

### Consent for publication

Not applicable

### Authors’ Contributions

M.P.M, N.K., E.T. and E.P. Writing -Original Draft, formal analysis, Visualization. M.P.M. and E.P. H.K., Z.M., M.H., S.D., J.B., G.J., Conceptualized, Methodology and funding acquisition. N.K., M.P.M: Project administration, Data Curation, MPM and EP: Supervision. M.P.M N.K., E.T., E.P., H.K., Z.M., M.H., S.D., J.B., G.J. J.C., F.W., E.V., S.N., S.N., L.M., S.P. :Investigation, Writing -Review & Editing.

## Acknowledgements

We would like to thank the members of our PPIE panels for their input and guidance with the SMILE intervention development process, and Anna Pethen for designing our intervention materials.

